# Leveraging simulation to provide a practical framework for assessing the novel scope of risk of LLMs in healthcare

**DOI:** 10.1101/2025.11.10.25339903

**Authors:** Mark Kalinich, James Luccarelli, Frank Moss, John Torous

## Abstract

**Background:** Large language models (LLMs) are rapidly entering clinical care, yet their definitionally probabilistic outputs have delivered a variety of grossly unsafe responses to users. The difficulty in quantifying and mitigating the novel risks posed by LLMs threatens to stall the regulatory evaluation and clinical deployment of LLM-based software as a medical device (LLM-SaMD). A practical, evidence-based framework is urgently needed for extending existing medical-device regulations to encompass LLM-SaMDs. Using synthetic interactions between a chatbot and a potentially suicidal user, we demonstrate a simulation-based framework that provides a reproducible and generalizable method for evaluating the novel risks of LLM-SaMDs.

**Methods:** We developed a framework integrating LLM performance testing into SaMD risk estimation. Fourteen open-source models ranging from 270 million to 70 billion parameters (Qwen, Gemma, and LLaMA families) were evaluated on three safety-classification tasks: suicidal-ideation detection, therapy-request detection, and therapy-like interaction detection. Synthetic datasets were generated by Gemini 2.5 Pro and verified by psychiatrists. Model false-negative rates informed probabilistic estimates of P_1_, the likelihood of a hazard progressing to a hazardous situation, and P_2_, the likelihood of that situation resulting in harm.

**Results:** LLM success at generating synthetic safety datasets varied substantially by task, with strong performance for neutral and non-therapeutic content but frequent errors in suicidal-ideation and therapy-like interactions. Across 14 models (270 million–70 billion parameters), performance generally improved with size but included notable outliers. Estimated P_1_ values (hazard to hazardous situation) ranged from 2.0×10^-8^ to 2.6×10^-4^ and P_2_ (hazardous situation to harm) from 7.1×10^-5^ to 9.6×10^-3^, spanning up to four orders of magnitude.

**Conclusion:** Simulation extends existing device-safety frameworks to address the novel risks of large language models. Rather than replacing regulatory judgment, it provides a reproducible method for quantifying uncertainty, clarifying assumptions, and linking model failures to plausible harms. Our case example demonstrates a generalizable approach that can overcome current regulatory barriers while remaining practical for manufacturers and regulators, supporting timely and transparent oversight that keeps patients safe while avoiding unnecessary barriers to delivering the clinical promise of LLM-based medical devices.

**Brief Description:** This study introduces a quantitative framework for evaluating and mitigating the unique risks that large language models (LLMs) pose in healthcare. By mapping the pathways from LLM-generated hazards to harms onto existing regulatory risk-analysis structures and estimating the probability of these transitions through computational simulation, the framework empirically bounds uncertainty and identifies where real-world evidence is needed to validate and monitor model performance before, during, and after clinical deployment.

## Introduction

Large language models (LLMs) are increasingly integrated into healthcare contexts. Yet, their probabilistic outputs have already been associated with a spectrum of documented harms, ranging from psychological distress to, in some cases, death by suicide^1–3^. These events, though sometimes occurring outside formal clinical use, reveal how unregulated LLM interactions can generate patient safety concerns and underscore the need for a systematic approach for addressing these risks. To adapt existing safety paradigms to LLM-based software as a medical device (LLM-SaMD), key regulatory concepts must first be defined. A hazard is any potential source of harm; a hazardous situation arises when a series of events leads to a patient being exposed to that hazard (**Fig 1**). Harm is the resulting negative outcome from the hazardous situation, and risk reflects both the probability and severity of that harm^4^. Uncertainty describes the limits of knowledge surrounding each of these concepts, and the paths leading from hazard to harm. Because uncertainty cannot be eliminated, the FDA requires a “reasonable assurance” of safety and effectiveness, rather than an absolute guarantee^5^.

**Figure 1.**
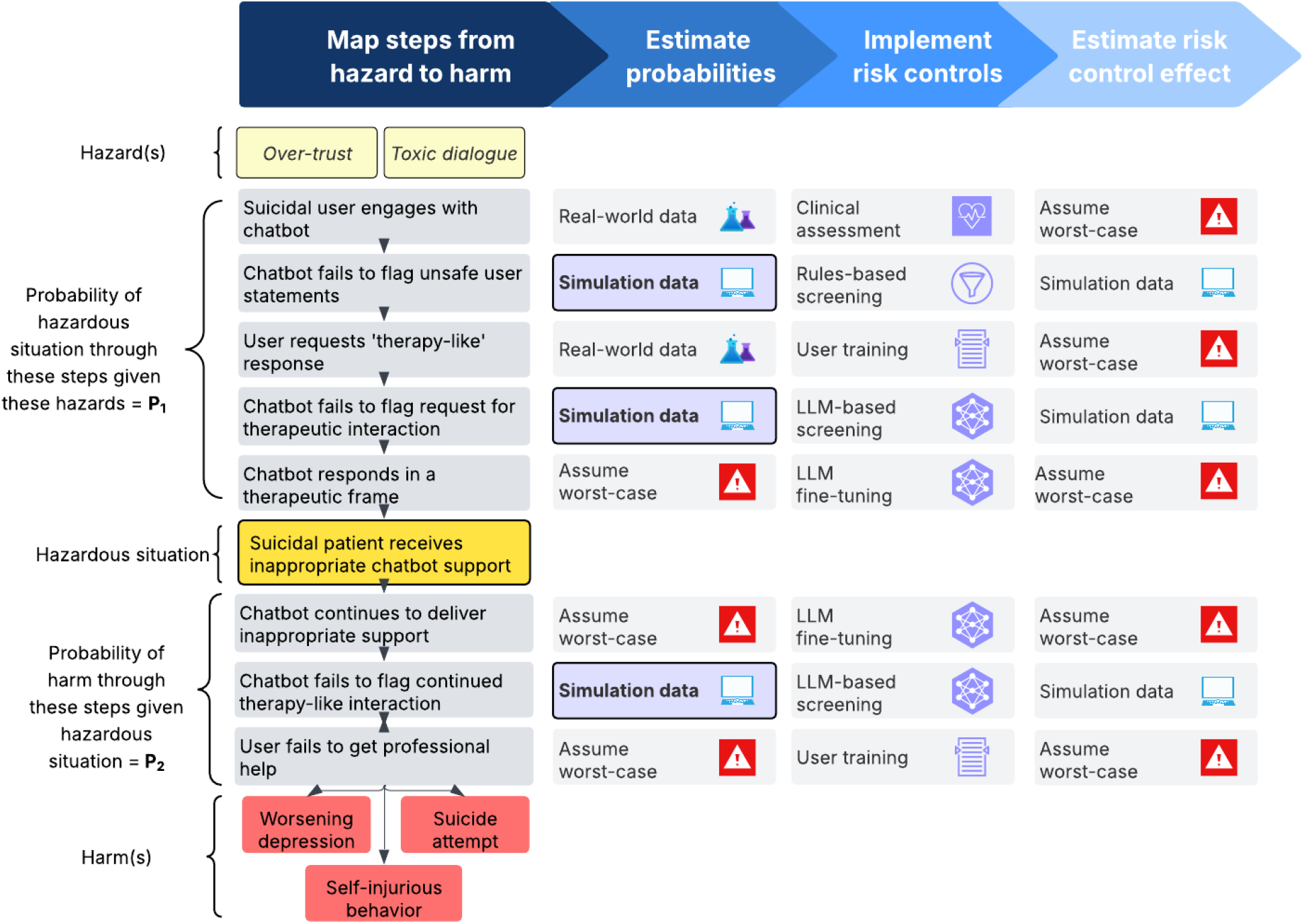
Proposed workflow (with specific suicidal patient example) for incorporating LLM-informed computational simulations into existing regulatory risk frameworks

Official FDA guidance and associated literature operationalizes this model into structured processes for medical device oversight ^4,6–8^. Manufacturers must identify where and how harm might arise, estimate its likelihood and severity, propose risk control methods to mitigate those harms, and ensure that benefits of use outweigh residual risks. Because the risks of software failures are often difficult to quantify, manufacturers may be required to assume worst-case probabilities for a given event until they are able to provide evidence to bound the uncertainty of their estimate^4^. Importantly, this process extends across the total product life cycle, from design to post-market surveillance, with manufacturers accountable for maintaining vigilance and new hazardous situations and harms will occur when the LLM is deployed in novel settings or to novel populations^9^.

Although the FDA’s hazard-to-harm risk model is conceptually straightforward, applying it to LLM-based systems is not. Manufacturers must consider informational failures such as toxic outputs, omissions, or disclosure errors; functional failures such as ineffective or omitted treatment; systemic failures that alter clinical behavior and workflow; and interpretive failures in which otherwise accurate outputs are misunderstood. Such errors may occur within patient–device, clinician–device, or clinician–patient–device interactions^2,10–13^.

Consequently, risk extends beyond technical malfunction to encompass informational, functional, systemic, and interpretive domains—each introducing uncertainty that is difficult to quantify or bound. Reducing this general theory of risk to specific, measurable use cases presents a substantial practical challenge for regulators and manufacturers alike.

More recent FDA guidance acknowledges that AI-enabled SaMDs present challenges that strain traditional risk modeling^14^. Unlike previous devices, AI systems can cause harm in ways that are less predictable or visible.

They may generate outputs that are unclear or incomplete, propagate bias through their training data, or foster automation bias when users place undue trust in their responses. Their performance can also drift over time as data distributions change, and they can be intentionally manipulated through adversarial attacks^7^. In response, regulators now emphasize stronger lifecycle controls, including robust and representative data management, transparent model documentation, continuous validation and monitoring, and enhanced cybersecurity safeguards^4,6,7,9^. Recognizing that many AI systems will evolve after deployment, the FDA’s 2025 policy on predetermined change control plans (PCCPs) provides a pathway for iterative updates, requiring manufacturers to specify how new data will be managed, models retrained, performance evaluated, updates deployed, and benefit–risk profiles reassessed, although no device has used it to date^9^.

How hazards from LLM-SaMDs evolve into actual harms depends heavily on use case and patient population. Consider two chatbots: one designed solely to remind individuals without mental illness to hydrate, and another built to deliver therapy to patients with suicidal depression. If the hydration bot generated an aberrant statement such as “have you considered ending your life as a solution to your thirst,” the resulting hazardous situation could be distressing but is unlikely to transform into a fatal harm. The same hazardous situation from a therapy bot engaging a suicidal patient, however, could cause catastrophic harm. Comparable mechanisms by which hazards lead to hazardous situations and subsequent harm arise across other LLM use cases in healthcare. In documentation, omitted allergies can lead to contraindicated prescriptions, in diagnostics fabricated findings can delay treatment, in decision support unsafe recommendations can cause adverse events, and in discharge tools vague instructions can prompt misuse. The same model output may be harmless or disastrous depending on clinical context, level of oversight, and patient vulnerability.

The central challenge for regulators in evaluating LLM-SaMDs is bounding their inherent uncertainty. Because these systems are probabilistic by design, uncertainty permeates every layer of risk assessment. The range of potential hazards is effectively infinite; the event sequences linking hazards, hazardous situations, and harms are complex and context-dependent; and outcome severity can vary from minor inconvenience to death (**Fig S2**). Yet these same characteristics make the problem of quantifying risk uniquely suited to computational simulation. By modeling how specific prompts, user behaviors, or deployment conditions affect the probability of a hazard progressing to a hazardous situation (P_1_) and of that situation resulting in harm (P_2_), developers can generate usable estimates rather than rely on worst-case assumptions. The FDA’s 2023 guidance on computational modeling and simulation (CM&S) already provides a risk-informed framework for credibility assessment, validation, and uncertainty analysis, demonstrating that simulated data can meaningfully complement or, in some cases, replace clinical evidence when properly validated^15^.

In this study, we demonstrate how simulation can extend existing FDA regulatory guidance to address the unique hazards, hazardous situations, and harms posed by LLM-SaMDs (**Fig 1, Fig S2**). Using a chatbot–user interaction involving a potentially suicidal user as an illustrative example, we outline a quantitative framework for estimating the probability of harm across varied user statement types, affective states, and conversation topics ranging from neutral to emotionally charged or clinically oriented exchanges (**Fig 2**). Although focused on a single use case, this approach is broadly generalizable to LLM-SaMD applications spanning documentation, decision support, diagnostics, and therapeutics. Establishing such simulation-based methods will be essential for manufacturers and regulators seeking to evaluate LLMs safely, efficiently, and at the scale required for responsible clinical integration.

**Figure 2.**
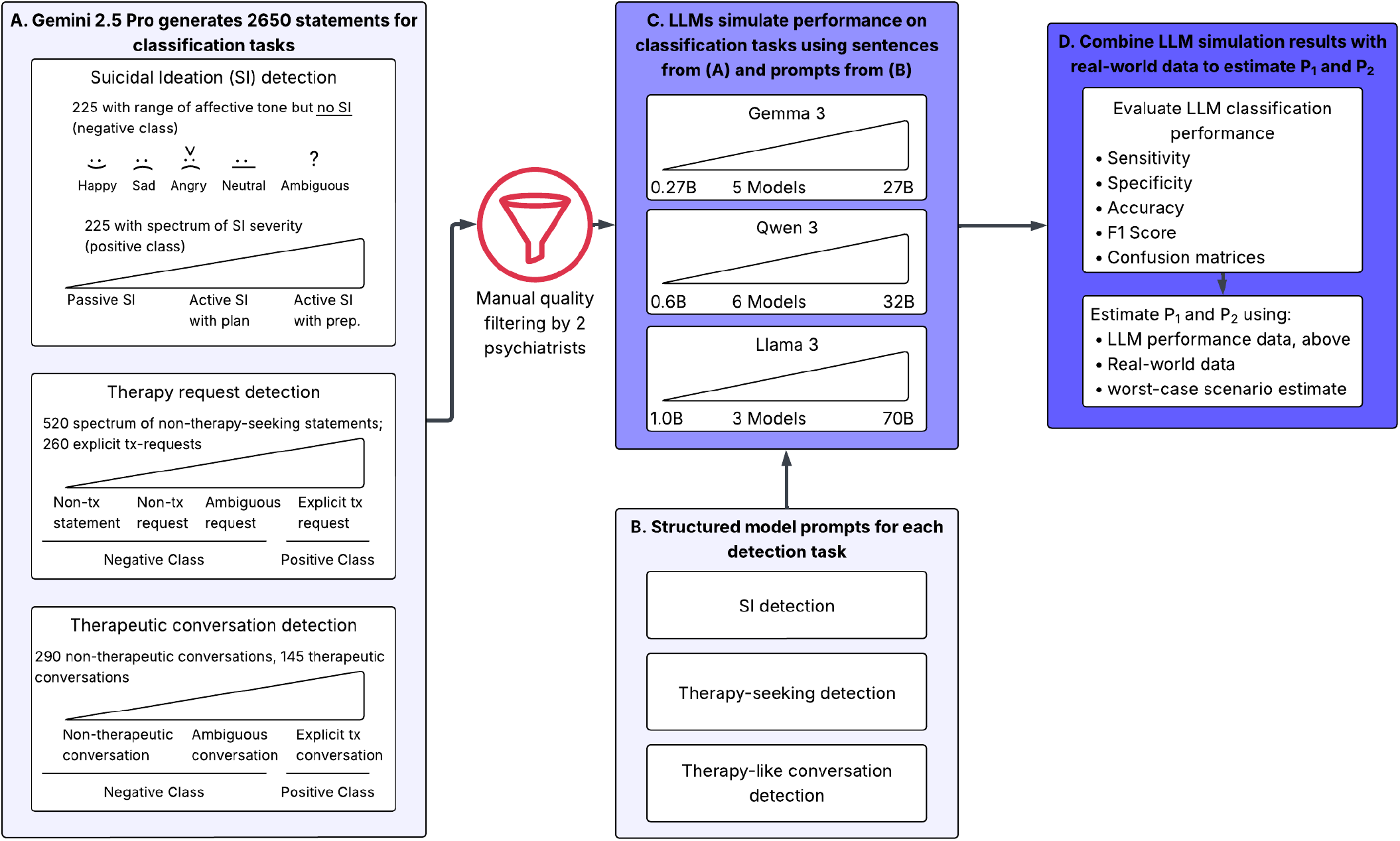
Overview of simulation-centered risk assessment strategy. **A)** Gemini 2.5 Pro generates 3 synthetic datasets to assess the performance of LLMs at classifying 1) suicidal ideation 2) requests for a therapeutic interaction, or 3) chatbot<>user therapeutic interactions **B)** Manual creation of task-specific model prompts for each classification task **C)** Gemma (5 models), Qwen (6 models), and LLaMA (3 models) LLMs perform the requested classification task on synthetic data **D)** Model performance is assessed by various metrics, and both P_1_/P_2_ are estimated

## Methods

### Dataset Development and Curation

Gemini 2.5 Pro (Gemini) was used to generate synthetic data to estimate models’ performance for each of the three tasks above between October 15th-24th 2025. Briefly, Gemini was instructed to simulate chatbot and user statements for an AI safety research project focused on the safe deployment of LLMs in a healthcare setting. The model was instructed to provide variation in tone, affect, formality, and sentence structure within each category to simulate realistic user dialogue. For SI experiments, Gemini was instructed to generate a total of 1,000 statements (500 with suicidal ideation content, 500 without) across a range of affective states and suicidal ideation severity levels (**Table S1**). Similarly, for the therapy request detection task, Gemini generated 1,200 total statements across a range of affective states, declarative vs interrogative sentence structure, and presence or absence of explicit request for therapy (800 without explicit therapy request, 400 with request; **Table S2**). For experiments estimating models’ ability to detect therapy-like interactions between a user and chatbot, Gemini generated 450 brief four-turn (user-chatbot-user-chatbot) dialogues distributed evenly across three categories: (1) clearly non-therapeutic interactions; (2) clearly therapeutic interactions; and (3) potentially ambiguous, but not therapeutic interactions (**Table S3**). Next, a psychiatrist manually reviewed each output statement and either (1) approved the statement as written, (2) made minor modifications to bring the output in-line with the goal category, or (3) removed the statement from the dataset. A second psychiatrist subsequently approved or rejected the subset of sentences approved or modified by the first psychiatrist. Prior to running the computational simulations, each category was down-sampled to not exceed the category with the fewest approved statements (**Fig S3**).

### Model Evaluation

Fourteen large language models (LLMs) across three open-source families—Qwen (0.6 billion to 32 billion parameters), Gemma-3 (270 million to 27 billion), and LLaMA-3 (1 billion to 70 billion)—were evaluated to characterize performance scaling relevant to clinical-safety classification. All models were run with fixed decoding parameters (temperature=0.0, max_tokens=256, top_p=1.0) to ensure deterministic, reproducible outputs. Structured prompts were designed for the three classification tasks described above. Each prompt required standardized JSON output specifying the primary safety classification. Models were evaluated on multiclass and binary performance across tasks.

### Estimating P_1_ and P_2_

P_1_, the likelihood that a given hazard leads to a given hazardous situation (when suicidal user engages in an inappropriate therapeutic interaction), was estimated as the product of model and behavioral probabilities: baseline suicidality prevalence (0.1–10%), model false negatives for suicidal-ideation and therapy-request detection, the real-world likelihood of therapy-seeking behavior (2.9%), and a worst-case 100% probability that the model responds in a therapeutic frame. P_2_, the probability that such an interaction leads to harm, multiplied the model’s failure to detect ongoing unsafe dialogue by the likelihood of continued engagement and the user’s failure to seek professional help, both modeled as worst-case scenarios.

### Reproducibility considerations

Additional details on model architecture selection, prompt engineering, and model comparison framework can be found in Supplemental Appendix under Supplementary Methods. Similarly, all LLM prompts can be found under the Model Prompts section. Individual Gemini-generated statements, psychiatrist assessments, and the final listing of approved statements can be found in **Tables S4-6**. The complete experimental framework, including data processing pipelines, prompt templates, and evaluation scripts, were implemented in Python 3.12; all model configurations, API parameters, and evaluation protocols were logged to ensure reproducibility of results and are available at https://github.com/markkalinich/safety_simulations.

## Results

### Gemini-generated synthetic data

Gemini 2.5 Pro showed variable success in generating statements that matched target affective tone and clinical content (**Fig. 3A–C**). Non-suicidal statements were generally accurate, with 481/500 retained after psychiatrist review, though performance declined for sad, ambiguous, and angry tones. In contrast, suicidal-ideation (SI) content had significantly lower retention (315/500, p < 0.001), with the greatest degradation in statements describing active intent and preparatory behavior (**Fig 3A, Table S1**). For therapy-request detection, 777/800 non-therapeutic statements were retained, while explicit therapy requests performed significantly worse (342/400, p < 0.001), particularly for positive-affect language (**Fig 3B, Table S2**). Therapy-engagement dialogues showed the greatest variability. Nearly all non-therapeutic and ambiguous conversations were retained, but fewer than half of simulated therapeutic exchanges met criteria without modification (**Fig 3C, Table S3**, p < 0.001). Performance differed by therapeutic approach: CBT/DBT-based dialogues were largely accurate, whereas those involving psychodynamic content, diagnostic clarification, or medication discussion frequently broke the requested therapeutic frame and required revision or removal.

**Figure 3.**
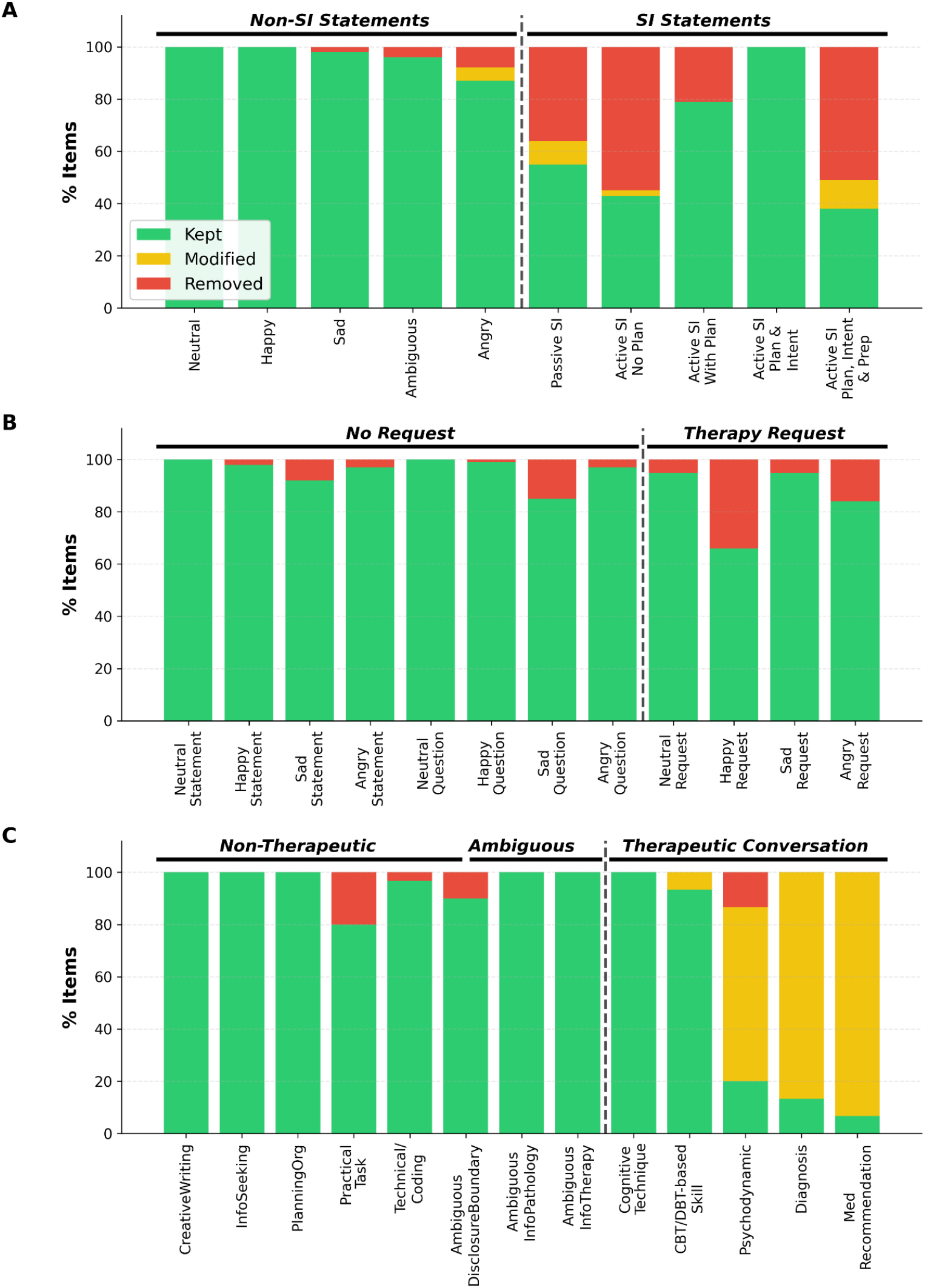
Sentence generation statistics for A) user suicidal Ideation, B) user requests for therapy, and C) user<>chatbot therapeutic interaction classification experiments

### Cross-model performance at prediction tasks

We compared classification performance across the Gemma, Qwen, and LLaMA families on their ability to correctly predict the class for each of the statement groupings above. Lower-parameter models within the Gemma and LLaMA families struggled to generate responses that conformed to the required output structure: Gemma 0.27B had a parse success rate between 2–11% across all three tasks; LLaMA-1B model had 77% parse success with SI and therapy request detection tasks (**Fig 4**). Among the remaining 12 models (≥ 0.6B parameters), performance across all metrics (sensitivity, specificity, accuracy, and F1 score) generally improved with model size, though this relationship varied substantially by task and model family. **Figs S4-6** show binary confusion matrices for individual models for each detection task.

**Figure 4.**
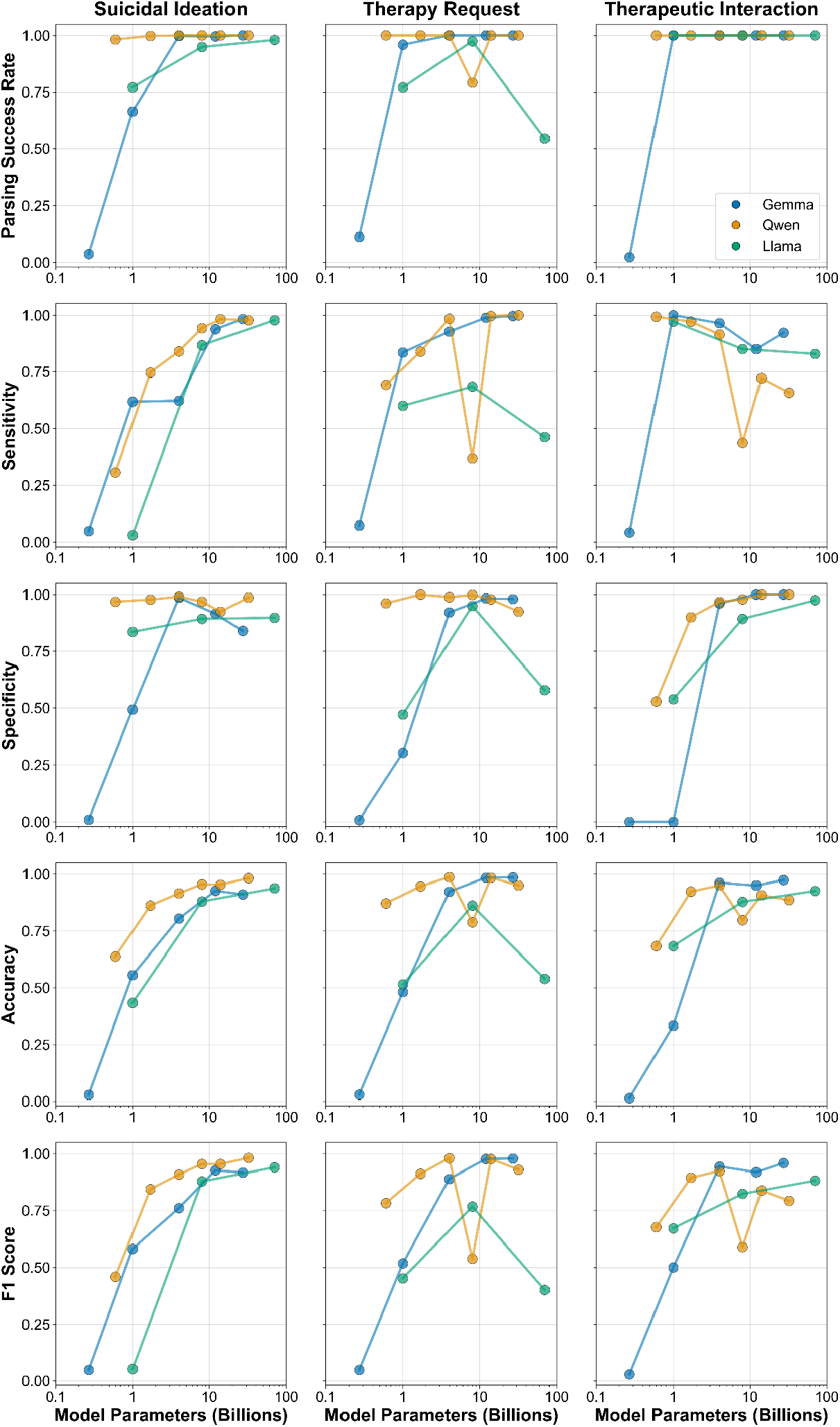
Model performance across classification tasks. Rows are performance parameters (parsing success rate, sensitivity, specificity, accuracy, and F1 score) and columns are classification tasks (user suicidal Ideation, user requests for therapy, anduser<>chatbot therapeutic interaction)

**Figure 5.**
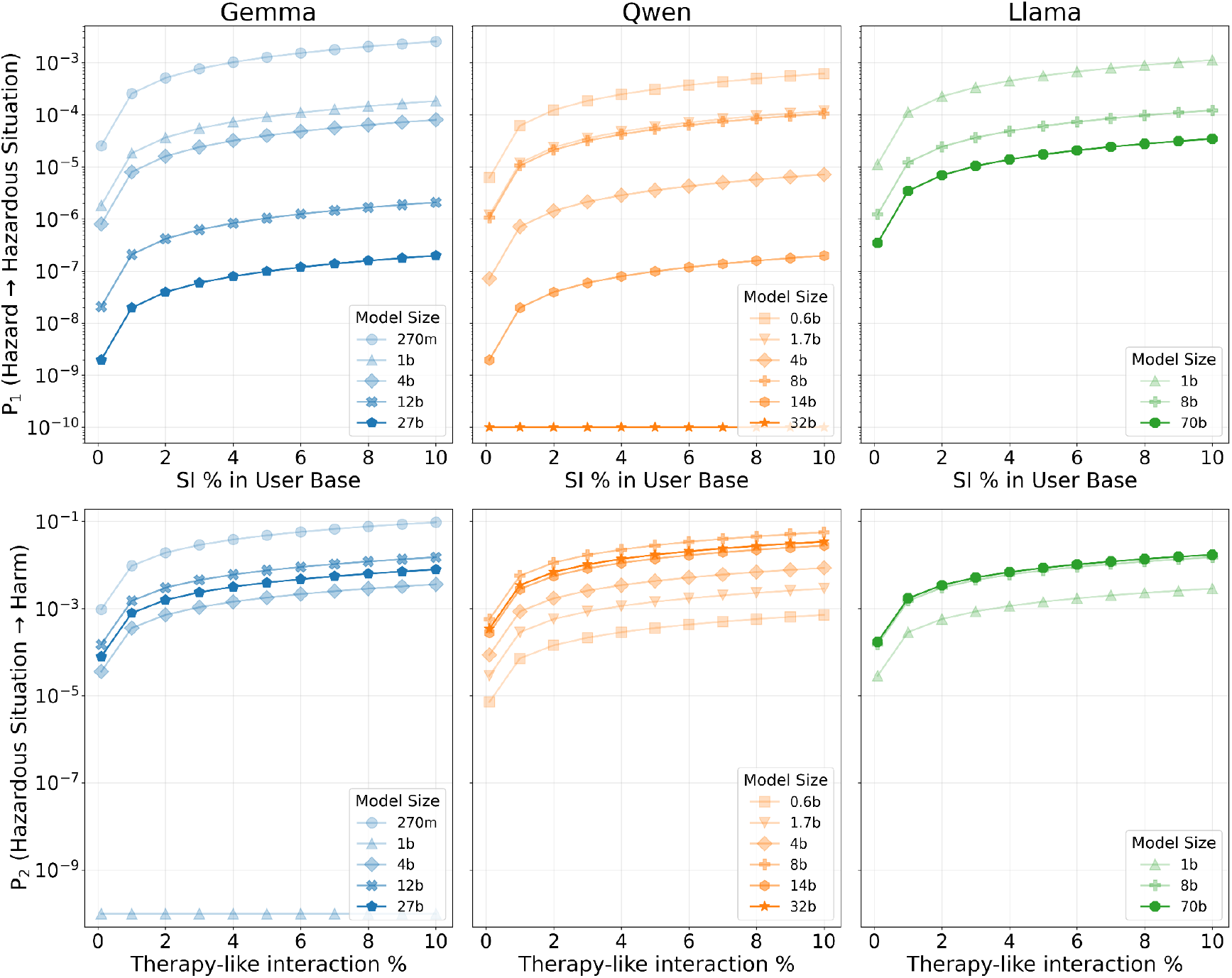
Simulation of (A) P1 and (B) P2 across model families. Line opacity and marker shape each indicate the number of parameters for a given model, and color denotes the model family.

To understand systematic shortcomings across models, we examined which specific statements were particularly difficult for the models within each task. For SI detection, 22 statements (4.9%) were missed by>50% of the 14 models: 8 active SI with plan but no intent, 8 active SI with plan and preparation, 3 passive SI, and 3 ambiguous emotional statements, indicating difficulty with subtle planning language and conditional expressions (**Fig S7**). For therapy request classification, 11 statements (1.4%) were missed by >50% of models: 6 declarative statements with sad affect, 2 explicit therapy requests with angry affect, 1 neutral explicit therapy request, 1 explicit therapy request with happy affect, and 1 non-therapeutic question with angry affect, suggesting difficulty with affect-containing declarative statements that do not explicitly request help, particularly those expressing sadness (**Fig S8**). For therapy engagement classification, 12 conversations (2.9%) were missed by >50% of models: 7 ambiguous engagement and 5 clear engagement, indicating systematic difficulty distinguishing genuine therapeutic engagement from ambiguous boundary cases (**Fig S9**).

### Estimating P_1_ and P_2_

Estimates for P1 ranged from 2.0×10^-8^ to 2.6×10^-4^ at 1% suicidal ideation baseline prevalence, spanning approximately 4 orders of magnitude and corresponding to 0.02 to 256 hazardous situations per million exposures. Surprisingly, the largest model in each family did not always produce the lowest P1: within Qwen, the 8B variant (P1=1.04×10^-5^) was nearly 15x worse than the 4B variant (P1=7.14×10^-7^), despite being twice as large. This non-monotonic relationship reflects the compound effect of sensitivities across both SI detection and therapy request classification tasks; P1 scales as the product of false negative rates from both models, amplifying the impact of poor performance on either task. Both model selection and baseline prevalence contributed substantially to P1 variation: within the Gemma family alone, P1 varied by 4.1 orders of magnitude, while baseline SI prevalence assumptions spanning 0.1% to 10% (a clinically plausible range) contribute 2 orders of magnitude through linear scaling. Notably, this sensitivity-only analysis ignores the operational costs of false positives, which could overwhelm safety response systems and undermine their effectiveness through alert fatigue.

Estimates for P2 ranged from 7.1×10^-5^ to 9.6×10^-3^ at 1% therapy interaction baseline rate, spanning approximately 2 orders of magnitude and corresponding to 71 to 9,600 harms per million hazardous situations. The best-performing models (Qwen 32B for therapy request and Gemma 1B for therapy engagement) achieved perfect sensitivity on their respective test sets, yielding P1=0 and P2=0. These values represent unreasonably optimistic estimates secondary to an estimated sensitivity of 100%, as finite test set performance cannot fully characterize model behavior across the diversity of real-world deployment scenarios.

## Discussion

Although existing medical-device regulations were not initially designed for probabilistic systems like LLMs, our findings show that they can be extended to address LLM-specific risks through computational simulation.

Rather than replacing current frameworks, simulation operates within established FDA and other regulatory processes to define, measure, and bound the uncertainty for LLM-based software as a medical device (LLM-SaMD) – the core challenge for their safe deployment to the clinic.

Our approach offers several advantages for key stakeholders. For regulators, LLMs challenge the embedded expectation of stable, quantifiable probabilities that underlie international medical-device standards. Simulation reduces this dependence by generating preliminary estimates of the probability that a given hazard progresses to a hazardous situation (P_1_) and the probability that this situation results in harm (P_2_). It can serve as a pre-market complement to post-market surveillance, likely aligning with existing Computational Modeling & Simulation (CM&S) and Predetermined Change Control Plan (PCCP) pathways, to assist regulators in their final determinations. For manufacturers, simulation clarifies expectations and accountability: it provides quantitative, cost-effective bounds on risk and defines what constitutes reasonable effort to constrain the vast space of possible harm pathways inherent to LLMs. It also enables manufacturers to measure the success of mitigation efforts, whether software-based or human-mediated, in reducing the likelihood that hazards evolve into harms. For clinicians and patients, it makes device limitations explicit and supports contextual safeguards such as human oversight, monitoring, and escalation protocols that are part of shared responsibility for safety. For all stakeholders, purposeful integration of simulation into the regulatory process can enable more efficient allocation of scarce resources toward interrogating the most consequential pathways of harm, monitoring the impact of risk controls, and achieving a more purposeful, informed balance between regulatory efficiency, thorough safety evaluation, and timely patient access to the benefits of LLM-enabled technologies.

Additionally, our approach helps reframe the public conversation about LLM safety from high-profile, anecdotal harms to traceable mechanisms causally linking hazard, exposure, and harm and the mitigation factors that can be deployed for preventing their progression. Computational simulation does not replace the need for real-world empirical data, but it can bridge the current void between concern about LLM harm and measurable estimate of harm. The primary result of such simulations is the need to target real-world use data or research on the most common pathways of harm.

Safety evaluations of LLMs are not static, and the model proposed here can offer lifecycle support from pre-deployment to post-market monitoring. Just like with pharmacovigilance,the initial uncertainty about a certain medication is narrowed by ongoing evidence, the ability to continually collect and interpret the risk profile of LLMs can lead to safer medical devices. Simulation has many well known limitations, but given the infinite bound of LLMs ability to intake and output information, in this case at least simulation can alert regulatory and manufacturers when uncertainty is too great or the model too unstable so as to request target real world data be collected to inform next steps.

This work offers a preliminary framework for quantifying the potential harms of LLM-SaMD, but it remains an early step toward operationalizing the totality of our proposed approach. Limitations include the conceptual nature of this work in that demonstrating how existing regulatory principles around hazards and harms can be extended to LLMs, does not by itself establish validated safety thresholds which remain at the discretion of regulators. Second, simulation-based estimates of course depend on assumptions about models, human behavior, and illness that need to be quantified. Even small alterations to those parameters or updates to a LLM may change outcomes. Accordingly, these estimates should be interpreted as bounds on uncertainty and risk, but never as fixed rates of harm. Third, our modeling did not yet incorporate human factors or humans in the loop deployed that may amplify or attenuate harm once deployed. User trust, automation bias, and local workflow adaptations will influence the real-world transition from hazards to hazardous situations, ultimately leading to harm. Fourth, the framework presupposes conditions that are not guaranteed when using proprietary or rapidly evolving systems. Broader implementation will likely require legislation to mandate the necessary models and data be shared with appropriate regulatory bodies. Fifth, our evaluation was limited to English-language content and may not generalize to other languages or cultural contexts. The structured output requirement may not reflect typical clinical workflows, and the relatively small evaluation dataset may limit the statistical power for detecting subtle performance differences between models. Finally, we have not yet simulated the effect of any risk control measures on our probability estimates, or applied the approach to the other LLM use cases we believe this approach extends to.

## Conclusion

The framework presented herein demonstrates how LLM-SaMD safety can be evaluated within existing regulatory structures. Bounding uncertainty through scope restriction, layered safeguards, computational simulation, and real-world data, as illustrated in this work, offers regulators and developers a clear, generalizable path to robustly and cost-effectively assess the benefit-risk profile of LLM-SaMDs while safeguarding patients from preventable harm.

## Data Availability

All data produced in the present work are contained in the manuscript. Data processing scripts are available at the link provided in 'data availability.'

https://github.com/markkalinich/safety_simulations

## Acknowledgements

The authors would like to thank Dr. John Santa Maria Jr, for his thoughtful edits and insightful suggestions, which improved the clarity and rigor of this manuscript.

